# TAS2R38 Taster Variants-Linked MGAM Expression in Alzheimer’s Disease: A Novel Target for Precision Drug Repurposing

**DOI:** 10.1101/2025.09.09.25334938

**Authors:** Claire W. Su, Kewei Chen, Teresa Wu, Eric M. Reiman, Qi Wang, Alzheimer’s Disease Neuroimaging Initiative (ADNI)

## Abstract

TAS2R38 is a taste receptor gene located on human chromosome 7 that influences sensitivity to bitter tastes and has been implicated in innate immunity, glucose level, and human longevity. However, its potential association with Alzheimer’s Disease (AD) has not been explored. Identifying such a genetic connection could support developing new drugs or repurposing existing ones for AD treatment. In this work, we examined the relationship between allele counts of TAS2R38 taster variants and AD risk using linear mixed-effects models, utilizing genetic, clinical, and biomarker data from the Alzheimer’s Disease Neuroimaging Initiative (ADNI). We investigated the potential molecular mechanisms of the association by identifying expression quantitative trait loci (eQTLs) using RNA-seq data from postmortem tissues across brain regions from the Religious Orders Study/Memory and Aging Project (ROSMAP). We evaluated whether FDA-approved drugs targeting the identified e-gene could reduce dementia risk using 1:1 propensity score-matched groups from longitudinal data in the National Alzheimer’s Coordinating Center (NACC) study, by comparing clinical dementia progression trends between the drug-taking and non-taking groups with linear mixed-effects models. Our results show that TAS2R38 supertasters were connected to a reduced AD risk with advancing age due to its association with various AD biomarkers (p < 0.001). eQTL analysis linked the nontaster allele to increased expression of the gene MGAM in AD-affected brain regions (p < 0.001). Furthermore, elevated MGAM expression correlated with more severe Tau burden (p < 0.05) and implicated in mitochondrial dysfunction in AD subjects. Notably, MGAM is a known drug target for diabetes mellitus. In NACC data, individuals taking MGAM-inhibiting drugs (acarbose and miglitol) showed slower clinical dementia rating progression (p < 0.01) in comparison with the non-taking group. This study is the first to report a genetic association between TAS2R38 and AD biomarkers. Our findings, validated in multiple cohorts/matching groups, suggest MGAM as a novel AD drug target with existing FDA-approved inhibitors and demonstrate the potential of TAS2R38 haplotypes to inform precision drug repurposing strategies for AD, which warrants further in-depth preclinical and clinical studies.

## 1. Introduction

TAS2R38 is a gene on human chromosome 7 that encodes a G-protein–coupled receptor involved in the perception of bitter tastes. It was first discovered when chemist Arthur Fox found varying individual responses to the taste of the chemicals phenylthiocarbamide (PTC) and 6-n-propylthiouracil (PROP), in which some individuals identified the taste of the chemical as bitter while others not. Studies identified three single nucleotide polymorphism (SNP) locations on the coding region of the gene that are responsible for this sensitivity of bitter taste compounds: rs10246939, rs1726866, and rs713598(Behrens et al., 2013). They result in three amino acid substitutions (A49P, A262V, I296V), with their haplotype combinations shown with the gene model in Figure 1A. The three sites are in strong linkage disequilibrium (R^2^ = 0.80−0.93, D’ = 0.997−0.998) with the PAV and AVI haplotypes predominant (> 90% frequency in a global population study) and more diversity in the African populations(Risso et al., 2016). The three most frequent genotypes are commonly referred to in the literature as supertaster (PAV/PAV), heterozygous taster (PAV/AVI), and nontaster (AVI/AVI) (Figure 1A).

**Figure 1:** TAS2R38 taster variants investigated for AD association, the mechanism and associated therapeutical potential. A). Gene model of TAS2R38 with the 3 relevant SNPs investigated in this work. The SNP IDs were simplified as g1-g3 throughout this manuscript. B). Project workflow to identify genetic association of TAS2R38 taster variants to AD and its linked MGAM gene expression as a novel drug repurposing target. Demographics for each cohort were reported in Table 1 and 2, as well as Supplementary Table 1-3. C). Flowchart for the procedures in selecting a propensity score-matched cohort of A10BF (acarbose or miglitol) ever users and never users in NACC data.

**Table 1:** Demographics for the subjects in this study from ADNI cohort (Okonkwo et al., 2025).

| taster group | super | hetero | non | other less<br>common<br>genotypes | all |
| --- | --- | --- | --- | --- | --- |
| longitudinal subjects | 436 | 922 | 616 | 194 | 2,168 |
| sex (M/F) | 232/204 | 481/441 | 329/287 | 118/76 | 1,160/1,008 |
| age at first visit (years) | 73.77±7.42 | 73.11±7.27 | 72.97±6.88 | 72.81±7.08 | 73.17±7.18 |
| education (years) | 16.18±2.62 | 16.02±2.82 | 16.16±2.62 | 16.02±2.93 | 16.09±2.74 |
| APOE4 allele count<br>(0/1/2) | 252/145/39 | 518/324/80 | 316/238/62 | 92/77/25 | 1,178/784/206 |

TAS2R38 has been implicated in innate immunity; it was found to be associated with susceptibility to diseases such as the early detection of pathogens in airways by using these receptors to distinguish helpful and harmful bacteria(Carey and Lee, 2019). It is also associated with longevity, with longer-lived groups having a higher supertaster ratio compared to groups with lower longevity(Melis et al., 2019). The nontasting variant (AVI) of the gene has also been shown to be a possible genetic risk for Parkinson’s Disease (PD)(Vascellari et al., 2020), and high postprandial glycemia(Gervis et al., 2025). However, its association with other neurodegenerative diseases such as AD has yet to be investigated. With the aging of global populations, AD has become the most devastating disease for the elderly with no effective treatment. Current treatment strategy primarily relies on symptomatic therapies like cholinesterase inhibitors and N-methyl-D-aspartate (NMDA) receptor antagonists, which offer modest cognitive benefits without altering disease progression. Recently, monoclonal antibodies targeting amyloid-β, such as

Lecanemab and Donanemab, have been approved as disease-modifying therapies for early-stage AD; although the drug conveys some benefits, it also comes with risks such as Amyloid-Related Imaging Abnormalities (ARIA)(Zimmer et al., 2025). Ongoing research focuses on tau pathology, neuroinflammation, and synaptic repair, with an increasing shift toward early intervention and biomarker-guided precision treatment(Zhang et al., 2024). Despite progress, AD remains incurable, and most therapies only modestly slow cognitive decline, warranting broader strategic approaches given the enormous complexity of its pathophysiology and associated comorbidities.

AD susceptibility has been linked to the composition of gut microbiota(Seo and Holtzman, 2024), innate immunity(Chen and Holtzman, 2022), or high glucose(Rawlings et al., 2017). Previous research has shown that TAS2R38 has mainly cytoplasmic and membranous expression in glandular tissues in the peripheral organs such as the tongue, respiratory tract, gut, and lungs(Douglas et al., 2019; Kobayashi et al., 2022), yet their variants could play a role in neurogenerative diseases via regulatory effects(Tong et al., 2025). By investigating the relationship between TAS2R38 haplotypes and AD, the underlying molecular mechanism could potentially lead to the discovery and implementation of new methods for precision treatment of AD, such as repurposed drugs that significantly reduce development time and cost. In this work, we investigated this TAS2R38 variants/AD relationship, beginning with the genetic, longitudinal clinical and biomarker data in the Alzheimer’s Disease Neuroimaging Initiative (ADNI). We reported a genetic association of the TAS2R38 taster variants with AD, specifically the lower risk of AD for the supertaster variants (PAV) with the advancement of aging and vice versa. From the genetic variants, we identified the linked eQTL gene MGAM, whose expression is significantly elevated in AD affected brain regions correlated with Braak staging. From causal inference test, the elevated MGAM expression was found to be implicated in mitochondrial dysfunction in AD. We additionally investigated the potential of inhibiting MGAM as a novel AD drug target since it is a target for type II diabetes (T2D) with FDA approved drugs. In propensity score matched groups of T2D patients, the group taking MGAM inhibiting drugs showed a significantly slow cognitive decline. The whole flowchart for this work is shown in Figure 1B. In each part of the work, the findings were validated in additional cohorts or by using different matching ratios in the propensity score matching process. These findings, including TAS2R38 genetic association to AD and the molecular mechanism underlying it, may enable new opportunities for precision drug repurposing in AD treatment.

## 2. Materials and methods

### 2.1 Genetic association of TAS2R38 taster variants to AD

Data were obtained from the ADNI database (adni.loni.usc.edu)(Okonkwo et al., 2025). The ADNI was launched in 2003 as a public-private partnership, led by Principal Investigator Michael W. Weiner, MD. The original goal of ADNI was to test whether serial magnetic resonance imaging (MRI), positron emission tomography (PET), other biological markers, and clinical and neuropsychological assessment can be combined to measure the progression of mild cognitive impairment (MCI) and early AD. The current goals include validating biomarkers for clinical trials, improving the generalizability of ADNI data by increasing diversity in the participant cohort, and to provide data concerning the diagnosis and progression of AD to the scientific community.

We downloaded the demographic, various longitudinal clinical assessment and biomarker data from ADNI data portal. Genetic data were obtained from the Alzheimer’s Disease Sequencing Project(Leung et al., 2025) (ADSP, https://adsp.niagads.org/). All genetic data were first converted into plink format using PLINK(Chang et al., 2015) for principal component analysis (PCA) of population structure. The plink genetic file on all the autosome chromosomes was used for PCA. We removed variant sites with missing data greater than 5%, minor allele frequency less than 5%, or significantly deviating from Hardy-Weinberg Equilibrium (HWE) (plink –geno 0.05 –maf 0.05 --hwe 1e-6). We further pruned the variants to exclude sites exhibiting high levels of linkage disequilibrium (LD), using the command “plink --indep-pairwise 50 5 0.2”. In addition, variants located in regions known to be under recent selection(Anderson et al., 2010) were removed from the dataset. PCA was performed in PLINK for the first 20 components, and based on the Scree plot, first five PCs were included in the subsequent association analysis.

We considered the alternative allele counts (dosages) of all three SNP sites to account for the diversity of the haplotypes and each variant’s contribution to the targets. The genotypes were extracted from vcf format using bcftools(Danecek et al., 2021) and converted into csv format. Cognitive tests and imaging biomarker data included clinical dementia rating (CDR, all six domains), PET-based amyloid loads (Centiloids), composite PET Tau scores (Braak staging converted from threshold-based tau-positivity in brain regions), and structural T1 MRI measurements from the ADSP Phenotype Harmonization Consortium (ADSP-PHC)(Hohman, 2023) harmonized datasets (release 3). The final dataset consists of 2,168 subjects with the demographic information reported in Table 1. All the data were read into R, where linear mixed effects (LME) models were built using the package “lmerTest”(Kuznetsova et al., 2017), comparing effects of the alternative allele counts (dosages) of the three SNPs (collectively referred to g1, g2, and g3 as shown in Figure 1A) to the longitudinal changes of the target variable with an interaction term for age, while controlling for several covariates such as sex, years of education, APOE4 allele count (the largest known genetic risk for AD), and the first five PCs, with the following equation in R programming language:

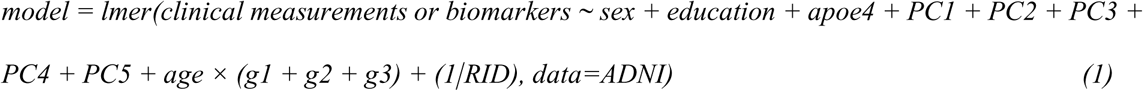

where RID is the unique subject ID.

We validated the results of this analysis in NACC (National Alzheimer’s Coordination Center: https://naccdata.org/requesting-data/nacc-data)(Beekly et al., 2007), with longitudinal data from over 42 current and former Alzheimer’s Disease research centers (ADRCs) across the US. Demographics, APOE4 allele count, and longitudinal clinical cognitive assessments (CDR) were obtained from Uniform Data Set (UDS) V3. Genetic data were obtained from the ADRC GWAS Datasets (ADC1-15) profiled by genotyping SNP array and harmonized by Alzheimer’s Disease Genetics Consortium (ADGC)(Naj et al., 2011). Samples closer than second-degree were removed in PLINK (--king-cutoff 0.0884). We only kept subjects with > 60 years of age at the first visit. Variant sites were pruned similarly as ADNI data for PCA. Based on the PCA, first 10 PCs were used in the LME models, where the targets were CDRs in six domains and the global and sum of boxes scores. The final dataset consists of 21,950 subjects with the demographic information reported in Supplementary Table 1.

### 2.2 eQTL identification and causal inference test

We performed the eQTL analysis based on the Accelerating Medicines Project for Alzheimer’s Disease (AMP-AD)(Hodes and Buckholtz, 2016) for the subjects within the Religious Orders Study/Memory and Aging Project (ROSMAP)(Bennett et al., 2018). All the demographic, clinical and pathological data of the ROSMAP cohort were obtained from the Rush Alzheimer’s Disease Center Research Resource Sharing Hub (https://www.radc.rush.edu/), upon approval of the data-usage agreement. Genetic data were from the variants called on whole genome sequencing (WGS) data from the AMP-AD data portal (https://www.synapse.org/Synapse:syn11707420). Additional genetic data were obtained from the AMP-AD diverse cohorts study (https://www.synapse.org/Synapse:syn51732523). The genetic data were merged and filtered similarly to what was done for the ADNI subjects, and PCA was performed. For gene expression data, we downloaded post-mortem gene expression data (RNA-seq) from three brain regions including dorsolateral prefrontal cortex (DLPFC), posterior cingulate cortex (PCC), and head of caudate nucleus (HCN) for the ROSMAP cohort from the AMP-AD data portal (https://www.synapse.org/Synapse:syn30821562)(Bennett et al., 2018). The gene expression data used was the filtered, normalized, and residualized counts to limit the effects of technical artifacts. After combining gene expression and genetic data, the final dataset consists of 947 unique individuals reported in Table 2.

**Table 2:**
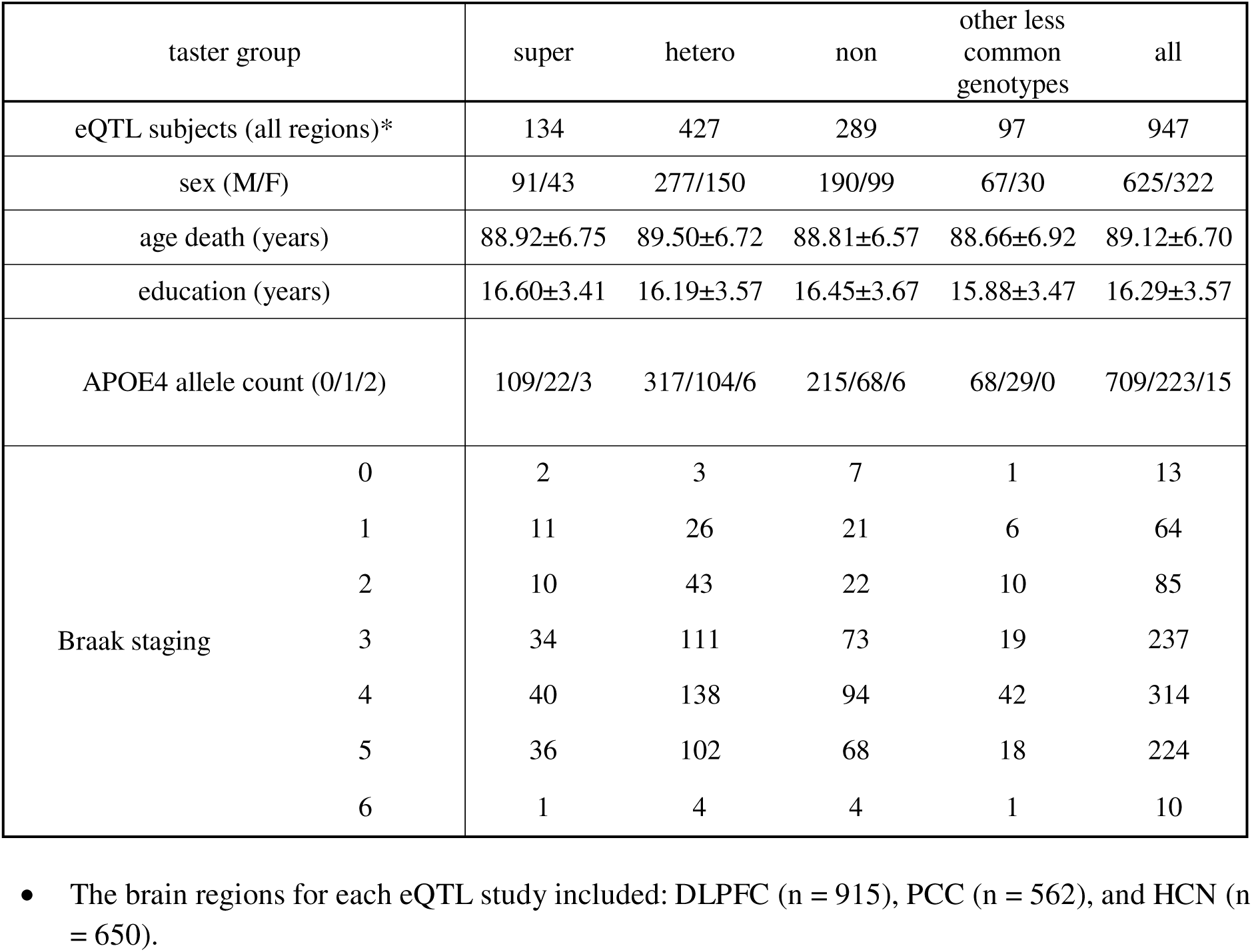
Demographics for the subjects from ROSMAP cohort (Bennett et al., 2018) for eQTL study (all regions).

We identified cis-eQTLs by linear regression of the expression of all the genes on chromosome 7 within 1Mb of the three SNPs (chr7: 140,792,804-142,973,545, 65 genes) with the alternative allele count of the three SNPs for each brain region using the following equation:

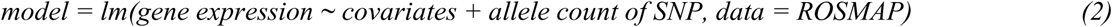

Covariates include technical covariates (RNA integrity number (RIN), post-mortem interval (PMI), sequencing batch (batch)), various biological covariates (age, sex, years of education, APOE4 allele count), and the first five PCs based on the PCA of the genetic data. Significant genes (e-genes) with FDR-corrected p value of less than 0.05 were then identified as eQTL of the SNP. The three SNPs were assessed individually for their corresponding e-genes. Semi-quantitative measurements of neuropathologies (Braak staging or CERAD score, Lewy body, TDP-43, and other vascular related neuropathologies) were added as covariates when they were additionally considered.

We carried out additional validations for the identified eQTL in two external cohorts from the AMP-AD study: the Mayo RNA-seq (MAYO) cohort(Allen et al., 2016) and the Mount Sinai Brain Bank (MSBB) cohort(Wang et al., 2018). The data were obtained from the same synapse entry as ROSMAP. The subject demographics are reported in Supplementary Tables 2 and 3 respectively. In MSBB, expression data were available for four brain regions: frontal pole (FP), inferior frontal gyrus (IFG), parahippocampus (PHG), and superior frontal gyrus (STG). For MAYO, two regions were profiled: temporal cortex (TCX) and cerebellum (CBE). Each region was assessed respectively for the eQTL. Neuropathology data for these two cohorts were downloaded from synapse entry syn27000096.

**Table 3:** Characteristics for matched group of A10BF drug users in NACC data (Beekly et al., 2007).

|  | User<br>(n = 38) | Non-user<br>(n = 38) | P<br>value | SMD |
| --- | --- | --- | --- | --- |
| **Demographic factors** |  |  |  |  |
| Age at first visit | 72.37±7.90 | 71.92±8.39 | 0.81 | 0.06 |
| Sex (M/F) | 20/18 | 20/18 | 1.00 | 0.00 |
| Education level (years) | 13.76±3.88 | 13.68±4.27 | 0.93 | 0.02 |
| Race (white/non-white) | 23/15 | 21/17 | 0.27 | 0.11 |
| Hispanic/Latino ethnicity | 11 | 9 | 0.79 | 0.12 |
| Family history of dementia | 13 | 4 | 0.03 | 0.59 |
| APOE ε4 (0/1/2/NA) | 18/4/2/14 | 18/4/2/14 | 1.00 | 0.00 |
| **Health behaviors** |  |  |  |  |
| Ever smoker | 19 | 18 | 1.00 | 0.05 |
| Alcohol abuse | 2 | 2 | 1.00 | 0.00 |
| Drug abuse | 1 | 0 | 1.00 | 0.23 |
| Obesity (BMI) | 30.43±8.51 | 29.25±9.09 | 0.54 | 0.15 |
| **Comorbidities** |  |  |  |  |
| Cardiovascular disease | 6 | 5 | 1.00 | 0.11 |
| Neurological diseases | 9 | 10 | 0.87 | -0.06 |
| Neuropsychiatric disorders | 16 | 17 | 1.00 | -0.05 |
| **Medication use** |  |  |  |  |
| Lipid □ lowering drugs | 27 | 28 | 1.00 | -0.06 |
| Anti □ hypertensive drugs | 31 | 34 | 0.52 | -0.22 |
| Non □ steroidal anti □ inflammatory medication | 16 | 17 | 1.00 | -0.05 |
| Antidepressants | 8 | 5 | 0.54 | 0.21 |
| Antipsychotic drugs | 0 | 1 | 1.00 | -0.23 |
| Anti □ Parkinson's drugs | 0 | 1 | 1.00 | -0.23 |
| Anxiolytic, sedative, or hypnotic agents | 4 | 9 | 0.22 | -0.35 |
| **Other diabetes drugs** |  |  |  |  |
| SGLT2 inhibitors (A10BK) | 0 | 3 | 0.24 | -0.41 |
| DPP □ 4 inhibitors (A10BH) | 10 | 5 | 0.25 | 0.34 |
| GLP □ 1RAs (A10BJ) | 3 | 6 | 0.48 | -0.25 |
| Sulfonylureas (A10BB) | 21 | 17 | 0.49 | 0.21 |
| Thiazolidinediones (A10BG) | 6 | 6 | 1.00 | 0.00 |
| Insulin (A10A) | 13 | 13 | 1.00 | 0.00 |
| Metformin (A10BA) | 22 | 22 | 1.00 | 0.00 |
| Other (A10BX) | 1 | 0 | 1.00 | 0.23 |

We performed causal inference test (CIT) to identify genes significantly affected by the strongest eQTL rs10246939 (g1) -MGAM using the RNAseq data of 502 AD subjects (defined as niareagansc < 3, i.e. NIA-Reagan diagnosis of AD(1997) high or intermediate) from DLPFC of ROSMAP. CIT has been well-described previously(Millstein et al., 2009). In short, it offers a hypothesis test for whether a molecule (in this case MGAM) is potentially mediating a causal association between a DNA locus (g1, i.e. rs10246939), and some other quantitative trait (such as the expression of genes correlated with MGAM and rs10246939). Causal relationships can be inferred from a chain of mathematic conditions, requiring that for a given trio of loci (L i.e. rs10246939), a potential causal mediator, (G, i.e. MGAM) and a quantitative trait (T, i.e. some other genes), the following conditions must be satisfied to establish that G is a causal mediator of the association between L and T:

a. L and G are associated
b. L and T are associated
c. L is associated with G, given T
d. L is independent of T, given G

We used the R software package “cit”(Millstein et al., 2016) to perform the causal inference test, calculating a false discovery rate using 1000 test permutations. Trios with a Q value (FDR) < 0.05 were classified as significant, and the associated T genes were considered downstream of MGAM. We investigated the significant gene hits for functional annotation and enrichment in knowledge databases such as KEGG(Kanehisa et al., 2023) pathways and Gene Ontology(Gene Ontology et al., 2023) terms through Metascape(Zhou et al., 2019).

### 2.3 Impact of MGAM inhibitor use on dementia risk and cognitive decline in diabetic subjects

To examine the effects of MGAM inhibiting drugs (acarbose or miglitol, Anatomical Therapeutic Chemical (ATC) code A10BF) on AD risk, we downloaded the comprehensive medical records including medication data from longitudinal assessments at NACC (National Alzheimer’s Coordination Center: https://naccdata.org/requesting-data/nacc-data)(Beekly et al., 2007), with data from over 42 current and former Alzheimer’s Disease research centers (ADRCs) across the US. Demographics, APOE4 allele count, longitudinal clinical cognitive assessments, comorbidities, and medication data were obtained from Uniform Data Set (UDS) V3.

The procedures used to create a cohort of 1:1 matched pair of ever users and never users of A10BF are shown in Figure 1C. We chose subjects with type II diabetes mellitus (T2D) based on diabetes diagnosis or use of antidiabetic medications at the first visit. Subjects with any dementia or PD diagnosis at the first visit were excluded. We matched the subjects who took MGAM inhibitors at any time in the follow-up period (ever-users) with another member of the never-user group with propensity score matching, using the R package “matchit”(Ho et al., 2011) by the following command:

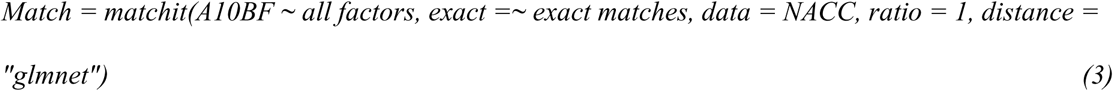

We considered the following factors in the matching: age at first visit, sex, race, years of education, APOE4 allele count, follow-up duration, total number of visits, and whether any other antidiabetic drugs taken in the duration of follow-ups, classified by their corresponding ATC subcategories (A10A (Insulin and analogs), A10BA (Biguanides), A10BB (Sulfonylureas), A10BG (Thiazolidinediones), A10BH (Dipeptidyl peptidase 4 (DPP-4) inhibitors), A10BJ (Glucagon-like peptide-1 (GLP-1) analogues), A10BK (Sodium-glucose co-transporter 2 (SGLT2) inhibitors), and A10BX (Other blood glucose lowering drugs)). Since it has been reported that A10BF could have joint effects with Metformin (A10BA) and Pioglitazone (A10BG) in reducing dementia incidences(Tseng, 2020), they were set as exact match together with Insulin (A10A), sex, and APOE4 allele count. To reduce overfitting in high-dimensional settings, we chose “glmnet” as the distance method to estimate propensity scores using L1/L2-regularized logistic regression(Friedman et al., 2010). Each individual in the user group was thus matched to the closest individual in the non-user group.

The final dataset contained 76 subjects in total, with 38 subjects taking MGAM inhibitors acarbose or miglitol, matched with non-users by a ratio of 1. Matching balance was evaluated with their subject characteristics reported in Table 3. Standard mean difference (SMD) was calculated for the two groups using the R package “tableone”(Yoshida and Bartel, 2022).

Using the time elapsed at first dementia diagnosis (DEMENT = 1), we performed survival analysis using the R package “survminer”(Kassambara et al., 2024) to examine the group difference in dementia susceptibility between ever-users and never users, and p-value was computed from the χ^2^ statistic in a log-rank test. Kaplan-Meier plots were created to present time to dementia diagnosis. To compare the drug’s effect on cognitive performances between the two groups, we built LME models to assess the group difference in multiple domains of CDR, by the following equation:

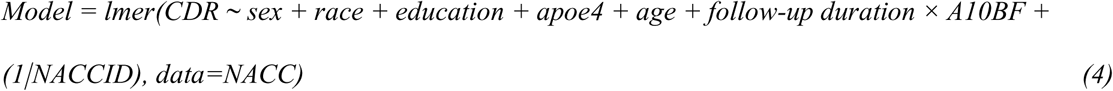

For robustness, we repeated the process with matching ratio set to 2 and 3 respectively.

### 2.4 Statistical analysis

Descriptive statistics for each study were reported for all subject characteristics using means and standard deviations for continuous variables and counts for categorical variables. Continuous variables were compared using a paired t-test between matched groups to check match balance. Categorical variables were compared using the Fisher’s exact test. P values from other models were obtained from each respective package. All the analyses were performed in R (v4.2.2) statistical language.

## 3. Results

### 3.1 TAS2R38 supertasters are associated with lower risk of AD with the advancement of age

From LME modeling of the longitudinal change in cognitive assessments and biomarkers in ADNI (n = 2,168), highly significant interactions (P < 0.001) were found between age and the alternative allele counts of each of the SNPs. Their contributions to the target variables are also significant (P < 0.001) yet in different directions (directed logP values and standardized coefficients reported in Figure 2, and raw effect size reported in Supplementary Table 4). This trend could be seen across different types of target variables, including multiple clinical and imaging biomarkers. The alternative alleles of g1 and g2 are found to have a negative effect on cognitive function in their interaction terms with age, while g3 has a positive effect; thus, with their comparable effect sizes, supertasters who have two copies of the alternative alleles at g1 and g3 (PAV haplotype, allele count combination 202) will have a lower negative effect on the cognitive performance thus slower cognitive decline when compared to nontasters with only two copies of the g2 alternative allele (AVI haplotype, allele count combination 020). Similar results were also found in imaging biomarkers from structural MRI and PET imaging data, especially for left lateral ventricle, a prominent biomarker for AD(Apostolova et al., 2012) (Figure 2A). The results were consistent although less significant in PET imaging for amyloid or tau burden (Supplementary Table 5, Supplementary Figure 1) due to smaller sample size (n = 1,767 for amyloid, n = 870 for Tau) with limited number of visits (mean = 2.2 for amyloid, 1.7 for Tau).

**Figure 2:** TAS2R38 supertasters are associated with a lower risk of AD with the advancement of age. A). Longitudinal changes in clinical assessment (CDRSUM) and imaging biomarker (left lateral ventricle volume), with fitted lines stratified by different taster groups. B). Heatmap of signed adjusted P-values showing correlations between predictor variables and target variables in the ADNI LME models. CDMEMORY: CDR memory score; CDORIENT: CDR orientation score; CDJUDGE: CDR judgement score; CDCOMMUN: CDR community affairs score; CDHOME: CDR home and hobbies score; CDCARE: CDR personal care score; CDGLOBAL: CDR global score; CDRSUM: sum of boxed for all CDR scores. Right lateral ventricle: right lateral ventricle volume; left lateral ventricle: left lateral ventricle volume. C). Forest plot of fixed effects from the LME model predicting CDRSUM, showing standardized coefficients with 95% confidence intervals (CI). Significant terms are colored in red.

The results were additionally validated using longitudinal data from NACC. There is more genetic diversity in the NACC study with larger sample size, yet the subjects possess more heterogeneous health conditions causing cognitive impairment, thus the effect size for each of the SNPs is smaller. Nevertheless, the supertaster allele was still found to be strongly associated with slower cognitive decline, surpassing the generally accepted genome-wide association study (GWAS) significance threshold (P < 5e-8, Supplementary Figure 2).

### 3.2 The TAS2R38 variants act as cis-eQTL of the gene MGAM to mediate AD risk

The e-genes from cis-eQTL for each of the alleles were reported in Table 4 respectively for all three regions (DLPFC, HCN, and PCC) from ROSMAP cohort. All three alleles are associated with multiple e-genes. In total, five genes were found to be significant across one or more brain regions in addition to DLPFC (WEE2-AS1, MGAM, TAS2R5, CLEC5A, and ENSG00000270157). Among them, WEE2-AS1 and MGAM show the strongest association with the g1 locus (adjusted P < 1E-10). When neuropathologies were added to the model, the expression of MGAM was also found to be significantly correlated with Braak staging (P = 0.028), where higher Braak staging associated with higher MGAM expression (Figure 3A). The association was only significant in DLPFC and not HCN or PCC (Table 4).

**Figure 3:** TAS2R38 taster variant mediates AD risk through the MGAM eQTL. A). MGAM expression in DLPFC split by rs10246939 (g1) alternative allele count and Braak staging from ROSMAP cohort. B). Causal inference network illustrating conditional relationships between locus rs10246939 (g1), MGAM expression, and downstream molecular networks implicated in AD. Red and blue arrows indicate down-and up-regulation respectively. C) Functional enrichment of the affected genes by the eQTL. n = 915, rs10246939 (g1) alternative allele count P = 5.6e-14, Braak staging P = 0.028.

**Table 4:**
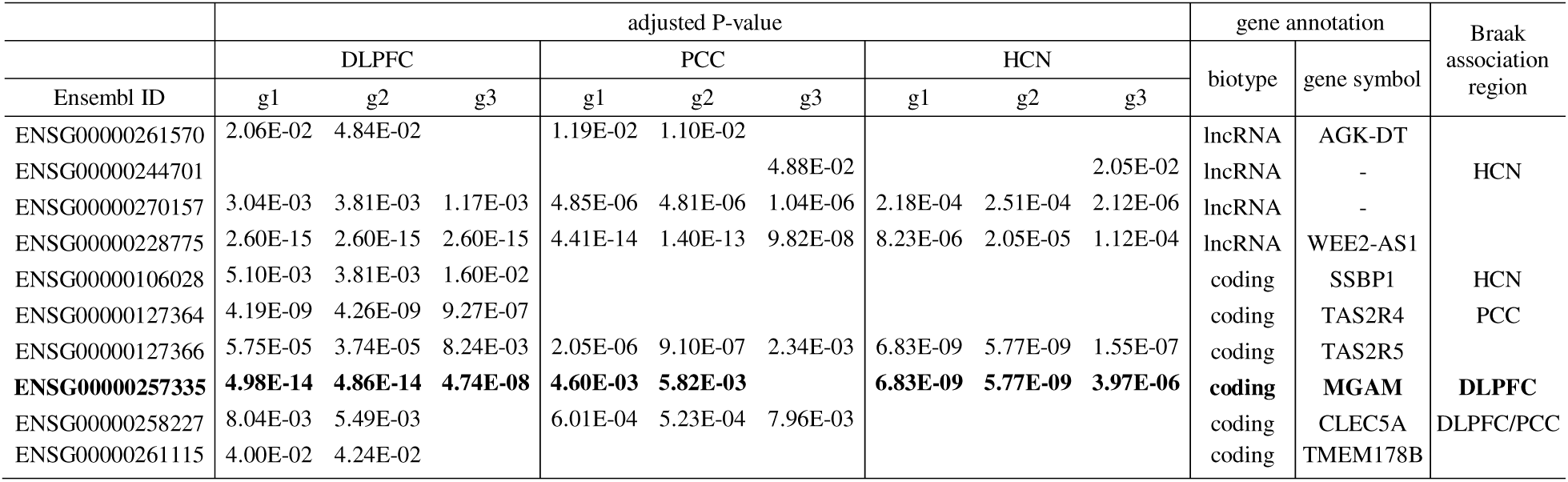
Significant eQTL genes of TAS2R38 variants across 3 brain regions in ROSMAP data. Only significant adjusted P values (< 0.05) are shown.

We further validated the eQTL relationship as well as associations of MGAM’s expression with neuropathology in different brain regions using the harmonized RNA-seq data from two additional cohorts of AMP-AD. Fewer e-genes were identified due to their smaller sample sizes. In MSBB, among the four brain regions (FP, IFG, STG, and PHG), eQTLs were identified only in IFG and STG, with MGAM as an e-gene in IFG. Interestingly, MGAM expression was also significantly associated with Braak staging in FP, PHG and STG (Supplementary Table 6, Supplementary Figure 3). In MAYO, g1 and g2 are in complete linkage disequilibrium, so their effects are exactly the same in opposite direction.

Their allele count and Braak staging were both significantly associated with MGAM expression in TCX, another AD vulnerable brain region (Supplementary Table 7, Supplementary Figure 4). Overall, alternative allele of the most significant locus g1 was found to be inversely correlated with MGAM expression, while neurofibril tangles were associated with increased MGAM expression in several AD affected brain regions, suggesting that lower MGAM expression confers lower AD risk for the supertasters and vice versa.

We further investigated the potential mechanistic role of TAS2R38 variants in the pathogenesis of AD through CIT analysis. Since the rs10246939 (g1) locus is most significantly associated with MGAM expression, we focused on the g1-MGAM network for the analysis. We identified 219 significantly affected downstream genes (FDR < 0.05, Supplementary Table 7), many of which encode for multiple units in the mitochondrial protein complex, such as VDAC3, VDAC4, CYC1, and UQCRC1 as well as 26S proteasome units such as PSMC4 and PSMD3. In addition, it was found that the expression of CHRNA7 (neuronal acetylcholine receptor subunit alpha-7), a major player of signal transmission at synapses, had been negatively impacted in the network. Molecular functional enrichments of the eQTL network revealed the disrupted gene modules to be implicated in AD (KEGG: hsa05010), learning or memory (GO: 0007611), and cognition (GO: 0050890) (Figure 3B, Supplementary Table 8). Some impacted genes are also associated with other neurodegenerative diseases (KEGG: hsa05022) such as PD (PSMC4, PSMD3, PRKN, CYC1, VDAC3, and UQCRC1) and Huntington’s disease (CYC1, PSMC4, PSMD3, UQCRC1, and VDAC3), indicating their roles in neurological functions (Figure 3C). The disrupted genes were negatively affected by increased MGAM expression, suggesting that higher MGAM expression drives protein degradation failure and mitochondrial damage, ultimately leading to synaptic loss.

### 3.3 MGAM inhibitors slow cognitive decline in T2D patients

Given MGAM’s role implicated in AD and the availability of FDA-approved inhibitors, we examined whether MGAM inhibiting drugs (A10BF) such as acarbose or miglitol could reduce AD risk or slow cognitive decline in T2D subjects by comparing matched groups with and without drug exposure in the NACC dataset, excluding individuals with Parkinson’s Disease (PD) for accurate propensity score matching.

We matched the individuals in the A10BF drug user group (ever-users) with those in the non-use group (never-users) with 1:1 propensity score matching. An evaluation of the matching balance is reported in Table 3. For the majority of the covariates such as age, sex, race, education, comorbidities, and commonly prescribed medication use, SMD between the two groups was found to be < 0.2, suggesting that the two groups were well matched in these areas. Some of the higher SMDs, such as those for the usage of certain medications, could be attributed to very small sample sizes of the observations in either group. However, the A10BF drug-taking group had a significantly larger (P = 0.026) portion with family history of dementia (either parent reported to have cognitive impairment).

We first compared the dementia-free probability in the two groups by setting the duration of dementia-free as the date from the first visit to the date where the first recorded dementia diagnosis was reported (i.e. first occurrence of DEMENT = 1). In the Kaplan-Meier plot comparing the groups with 1:1 matching, a borderline significant trend (P = 0.059) can be seen, with the non-users having an earlier dementia diagnosis rate or a higher AD susceptibility than the drug user group (Figure 4A). This suggests that the drug could play a role in decreasing dementia risk in T2D patients.

**Figure 4:** MGAM inhibitors slow down cognitive decline in T2D patients. A) Kaplan-Meier curve comparing dementia-free probability between ever users and never users of MGAM inhibitors in PSM groups at ratio=1. n = 38 x 2, P = 0.059. B) Longitudinal changes in clinical assessment (CDRSUM) with fitted lines stratified by drug user groups in A). Group difference P < 0.001. Inset is standardized coefficients for the LME model. Significant terms are colored in red.

Since dementia is a progressive neurodegenerative condition rather than a binary state, we further examined the drugs’ effects on cognitive performance by comparing longitudinal CDR measurements between the ever-user and never-user groups. In addition to overall CDR deterioration over time (P < 0.001 for follow-up duration), we observed a significant group difference in CDR change (interaction between follow-up duration and A10BF status) across all six domains (P < 0.05 for each domain and P < 0.001 for CDRSUM; Figure 4B, Supplementary Table 9), demonstrating a strong association between drug use and cognitive outcomes. The drug taking group showed a significantly slower cognitive decline, especially for orientation (P < 0.001), community affairs (P < 0.001), personal care (P < 0.01), and home and hobbies (P < 0.01). These observations still held when we increased the matching ratio to 2 and 3 respectively (Supplementary Figure 5, Supplementary Table 9), strengthening the possibility that A10BF could help ameliorate cognitive decline in T2D subjects.

## 4. Discussion

We reported for the first time the genetic association of TAS2R38 taster variants with AD, with the supertasters (PAV haplotype) exhibiting a reduced risk of AD with the advancement of age and vice versa. The results were validated in multiple cohorts by various clinical cognitive assessments and biomarkers.

Notably, the association has not been reported in previous case-control GWAS results (Kunkle et al., 2019; Bellenguez et al., 2022), likely for the reasons:

1. The effect is only significant in older age, requiring statistical models that include an interaction term with age.
2. The observed association arises from the combined effect of multiple haplotypes. Because each of the three SNP alleles contributes with different effect sizes and directions, their full impact is only revealed when analyzed together in diverse cohorts.

When the rare haplotypes (other than super-, heterozygous-, or non-tasters) are excluded in the model, we can still identify the supertaster allele (PAV) associated with slower cognitive decline, albeit less significantly (n = 1,974; P = 5.47e-4 for allele and 1.42e-4 for age*allele when target = CDRSB). This is expected, as g3 is not as close in genomic distance as the other two variants (g1/g2, which are within 100 bp), so the linkage is weaker and it cannot tag the haplotype as effectively. Only when they were modeled jointly could their individual effects be fully revealed. If the model is built without an interaction term with age (as it has been for quantitative traits in conventional GWAS), the association is even weaker (P = 0.01-0.04 for the three SNPs respectively when tested separately), which is comparable to those observed in case-control GWAS from large biobanks such as FinnGen(Kurki et al., 2023) with the same effect direction. This underscores that, for a complex genetic makeup, individual variants need to be modeled jointly(Manolio et al., 2009) to fully reveal their different effects, and it highlights the importance of modeling joint allele effects in the context of age(Winkler et al., 2024) when investigating the genetic architecture of neurodegenerative diseases, which manifest over a long time course. Notably, these effects are larger in the AD patients (such as those from ADNI) than dementia in general (such as those from the heterogeneous cohort NACC), suggesting the specific roles they play in AD.

We further investigated the molecular mechanism underlying the association. Since the expression of TAS2R38 in brain tissues is low to negligible in subjects in public databases such as GTEx(Consortium, 2013) and AD-related datasets such as those from AMP-AD, the gene is unlikely to directly modulate brain functions. We postulated that the association between the taster genotype and AD is mediated by different TAS2R38 allele counts affecting the expression of nearby AD-implicated genes through a cis-eQTL mechanism. Indeed, the expression of the gene MGAM (maltase-glucoamylase) was found to be significantly correlated with TAS2R38 taster variants across several AD affected brain regions, supported by data from three independent cohorts, even those with smaller sample sizes such as MAYO. In addition, its expression is also correlated with neurofibril tangles (Braak staging) in many brain regions such as DLPFC, TCX, and PHG. Previously higher expression of MGAM has been identified to have causal effects on the increased risk of AD through two-sample Mendelian randomization analysis(Meng et al., 2022). As a gene coding for an enzyme that breaks down glucose, it has been a target for diabetes treatment with the FDA approved drugs such as acarbose and miglitol. AD has been suggested as a metabolic disease(Nguyen et al., 2020), with lots of ongoing studies researching the association between diabetes and dementia(Tseng, 2020). Therefore, MGAM is prioritized in this study for in depth investigation as a mediator of the association between TAS2R38 variants and AD.

Through CIT analysis, we identified 15 genes impacted by the rs10246939(g1)/MGAM eQTL implicated in pathways central to AD and cognition. The expression of most of these genes is negatively correlated with increased MGAM expression. The location and known functions of these genes show a clear link to mitochondrial dysfunction, proteasome impairment and synapse failure as a person ages. These processes are critical aspects of neuronal stress response and are recognized as key causes of AD. These findings suggest that inhibiting MGAM could be a promising therapeutic strategy for AD.

We further examined if existing inhibitors targeting MGAM (A10BF) could help ameliorate cognitive decline in T2D patients, leveraging the comprehensive longitudinal cognitive evaluations at NACC coupled with their medication data. T2D is a long-suggested risk factor for AD. While many T2D drugs have been investigated for their potential to influence dementia risk in T2D patients, the results have been mixed and contradictory(Kuate Defo et al., 2024). To address this, we matched the ever-user group of A10BF with the never-users based on the propensity score including their specific T2D medication categories, as well as other established risk factors such as demographics and APOE4 allele count. The matching groups show a relatively balanced distribution of other diabetic drug exposure, ensuring a fair comparison of A10BF’s effects.

We observed that A10BF use was associated with a suggestive lower risk of developing dementia in the matched groups in survival analysis. The lack of definite statistical significance can be attributed to the relatively small sample size, since acarbose or miglitol are not widely used as T2D treatment in western countries. In addition, dementia is not a binary state but more a progressive condition, so the exact time of onset cannot be accurately determined. The drugs’ effects are better demonstrated in longitudinal cognitive performance, where we observed a clear significant difference in CDR between the two groups, with the ever-users showing slower cognitive decline. Studies have suggested that MGAM inhibitors are beneficial to cognitive function in rodent models(Manandhar et al., 2025; Sonsalla et al., 2025) and reducing dementia risk in population-based retrospective cohort study(Tseng, 2020), yet our work provided further evidence and revealed an underlying genetic architecture for MGAM as a novel drug target for AD.

T2D and AD share several pathophysiological mechanisms, including impaired glucose metabolism, insulin resistance, mitochondrial dysfunction, chronic inflammation, and vascular injury. These overlaps have led to substantial interest in repurposing T2D medications for AD prevention or treatment. Several drug classes, especially the GLP-1RA class (A10BJ) have been evaluated in epidemiological, preclinical, and clinical studies, yet so far no concrete outcome has been reported. This underscores the complexity of the disease mechanism and gene-environment interplay and highlights the need for targeted mechanistic and clinical investigations. Previous work demonstrated that TAS2R38 modulates postprandial glucose levels by triggering dose-dependent secretions of GLP-1(Wang et al., 2025). The effect of its variants on glucose level is reported to be independent of diet and lifestyle(Gervis et al., 2025), suggests a mechanism beyond dietary preference. In our investigation from NACC data, the two treatment groups didn’t show much difference in health behaviors either, supporting a role A10BF plays in cognition instead of differential dietary habits by taster sensitivity since we didn’t consider TAS2R38 genotypes in the matching. Current FDA-approved MGAM inhibitors offer poor (acarbose) to poor-moderate (miglitol) brain permeability, so their pharmacological effects on cognitive functions could be limited by their inaccessibility to the target in the brain(Ogura and Yamaguchi, 2022). Further optimization of their pharmacokinetic and pharmacodynamic properties is needed to evaluate their full potential for repurposing as treatments for AD.

Our study has several limitations. One limitation is that the sample size was limited in the investigation of the effects of MGAM-targeting drugs on dementia risk and cognitive performance due to the rare use of A10BF for T2D treatment in the US. From an observational study such as NACC, complete data on drug dosage, cumulative exposure, duration of therapy, or adherence are not available, so it is impossible to infer a pharmacologic relationship between drug intensity and cognitive outcome. To boost power, we have included the “ever-users” of self-reported A10BF use in the whole follow-up period, with a mean (visit number, or NACCVNUM) = 2 for first report use and mean exposure = 2 (visits). If we just include the users who reported taking A10BF at the baseline visit (NACCVNUM = 1), the user group would include only 24 subjects. A comparable 1:n (n =1 to 3) matching with non-users still yielded consistent, significant group differences in follow-up CDR measurements (Supplementary Table 10), supporting the robustness of the result. Another limitation is that TAS2R38 genotypes are not fully available for the NACC subjects, so we could not include the taster categories in the propensity score for precision matching, thus lacking a detailed analysis of the drugs’ effect on different taster genetic makeups. We postulate that genetically predisposed differences in MGAM expression levels in the brain due to TAS2R38 genetic variants confer differential dementia risk, offering the opportunity for precision drug repurposing. In the future, we anticipate leveraging genetic and detailed medical records data from large biobanks such as All of Us(2019) and UK Biobank(Sudlow et al., 2015) for in-depth research in this direction to enable personalized AD treatment. Ultimately, more preclinical and clinical studies are warranted to further investigate the potentials of MGAM-inhibiting drugs as an alternative or supplementary treatment option for AD.

## Data Availability

This work utilized all public datasets in the analyses with data use application required. ADNI data are available at https://adni.loni.usc.edu/. Datasets for eQTL analysis are available on AMP-AD knowledge portal (https://adknowledgeportal.synapse.org/) with synapse accession ID included in the main text. All the clinical and pathological data for the ROSMAP cohort were obtained from the Rush Alzheimer's Disease Center Research Resource Sharing Hub (https://www.radc.rush.edu/). NACC data are available at https://naccdata.org/. Additional genetic data from ADSP are available at https://adsp.niagads.org/.

## Acknowledgements

Data used in preparation of this article were obtained from the Alzheimer’s Disease Neuroimaging Initiative (ADNI) database (adni.loni.usc.edu). As such, the investigators within the ADNI contributed to the design and implementation of ADNI and/or provided data but did not participate in analysis or writing of this report. A complete listing of ADNI investigators can be found at: http://adni.loni.usc.edu/wp-content/uploads/how_to_apply/ADNI_Acknowledgement_List.pdf. Data collection and sharing for this project was funded by the Alzheimer’s Disease Neuroimaging Initiative (ADNI) (National Institutes of Health Grant U01 AG024904) and DOD ADNI (Department of Defense award number W81XWH-12-2-0012). ADNI is funded by the National Institute on Aging, the National Institute of Biomedical Imaging and Bioengineering, and through generous contributions from the following: AbbVie, Alzheimer’s Association; Alzheimer’s Drug Discovery Foundation; Araclon Biotech; BioClinica, Inc.; Biogen; Bristol-Myers Squibb Company; CereSpir, Inc.; Eisai Inc.; Elan Pharmaceuticals, Inc.; Eli Lilly and Company; EuroImmun; F. Hoffmann-La Roche Ltd and its affiliated company Genentech, Inc.; Fujirebio; GE Healthcare; IXICO Ltd.; Janssen Alzheimer Immunotherapy Research & Development, LLC.; Johnson & Johnson Pharmaceutical Research & Development LLC.; Lumosity; Lundbeck; Merck & Co., Inc.; Meso Scale Diagnostics, LLC.; NeuroRx Research; Neurotrack Technologies; Novartis Pharmaceuticals Corporation; Pfizer Inc.; Piramal Imaging; Servier; Takeda Pharmaceutical Company; and Transition Therapeutics. The Canadian Institutes of Health Research is providing funds to support ADNI clinical sites in Canada. Private sector contributions are facilitated by the Foundation for the National Institutes of Health (www.fnih.org). The grantee organization is the Northern California Institute for Research and Education, and the study is coordinated by the Alzheimer’s Disease Cooperative Study at the University of California, San Diego. ADNI data are disseminated by the Laboratory for NeuroImaging at the University of Southern California.

The results published here are in part based on data obtained from the AD Knowledge Portal (https://adknowledgeportal.org). The RNA-seq Harmonization Study was supported by the NIA grants U01AG046152, U01AG046170, U01AG046139 and U24AG061340. Study data in ROSMAP cohort were provided by the Rush Alzheimer’s Disease Center, Rush University Medical Center, Chicago, IL, USA. Data collection was supported through funding by NIA grants P30AG10161 (ROS), R01AG15819 (ROSMAP; genomics and RNA-seq), R01AG17917 (MAP), R01AG36836 (RNA-seq), the Illinois Department of Public Health and the Translational Genomics Research Institute. Additional phenotypic data were requested at https://www.radc.rush.edu. The data for MSBB cohort were generated from post-mortem brain tissue collected through the Mount Sinai VA Medical Center Brain Bank and were provided by Dr Eric Schadt from Mount Sinai School of Medicine. The MSBB study was led by Dr Nilufer Ertekin-Taner and Dr Steven G. Younkin, Mayo Clinic, Jacksonville, FL, USA, using samples from the Mayo Clinic Study of Aging, the Mayo Clinic Alzheimer’s Disease Research Center and the Mayo Clinic Brain Bank. Data collection was supported through funding by NIA grants P50AG016574, R01AG032990, U01AG046139, R01AG018023, U01AG006576, U01AG006786, R01AG025711, R01AG017216, R01AG003949, NINDS grant R01NS080820, CurePSP Foundation and support from Mayo Foundation. Study data include samples collected through the Sun Health Research Institute Brain and Body Donation Program of Sun City, Arizona. The Brain and Body Donation Program is supported by the NINDS (U24NS072026 National Brain and Tissue Resource for Parkinson’s Disease and Related Disorders), the NIA (P30AG19610 Arizona Alzheimer’s Disease Core Center), the Arizona Department of Health Services (contract 211002, Arizona Alzheimer’s Research Center), the Arizona Biomedical Research Commission (contracts 4001, 0011, 05-901 and 1001 to the Arizona Parkinson’s Disease Consortium) and the Michael J. Fox Foundation for Parkinson’s Research.

The NACC database is funded by NIA/NIH Grant U24 AG072122. NACC data are contributed by the NIA-funded ADRCs: P30 AG062429 (PI James Brewer, MD, PhD), P30 AG066468 (PI Oscar Lopez, MD), P30 AG062421 (PI Bradley Hyman, MD, PhD), P30 AG066509 (PI Thomas Grabowski, MD), P30 AG066514 (PI Mary Sano, PhD), P30 AG066530 (PI Helena Chui, MD), P30 AG066507 (PI Marilyn Albert, PhD), P30 AG066444 (PI David Holtzman, MD), P30 AG066518 (PI Lisa Silbert, MD, MCR), P30 AG066512 (PI Thomas Wisniewski, MD), P30 AG066462 (PI Scott Small, MD), P30 AG072979 (PI David Wolk, MD), P30 AG072972 (PI Charles DeCarli, MD), P30 AG072976 (PI Andrew Saykin, PsyD), P30 AG072975 (PI Julie A. Schneider, MD, MS), P30 AG072978 (PI Ann McKee, MD), P30 AG072977 (PI Robert Vassar, PhD), P30 AG066519 (PI Frank LaFerla, PhD), P30 AG062677 (PI Ronald Petersen, MD, PhD), P30 AG079280 (PI Jessica Langbaum, PhD), P30 AG062422 (PI Gil Rabinovici, MD), P30 AG066511 (PI Allan Levey, MD, PhD), P30 AG072946 (PI Linda Van Eldik, PhD), P30 AG062715 (PI Sanjay Asthana, MD, FRCP), P30 AG072973 (PI Russell Swerdlow, MD), P30 AG066506 (PI Glenn Smith, PhD, ABPP), P30 AG066508 (PI Stephen Strittmatter, MD, PhD), P30 AG066515 (PI Victor Henderson, MD, MS), P30 AG072947 (PI Suzanne Craft, PhD), P30 AG072931 (PI Henry Paulson, MD, PhD), P30 AG066546 (PI Sudha Seshadri, MD), P30 AG086401 (PI Erik Roberson, MD, PhD), P30 AG086404 (PI Gary Rosenberg, MD), P20 AG068082 (PI Angela Jefferson, PhD), P30 AG072958 (PI Heather Whitson, MD), P30 AG072959 (PI James Leverenz, MD).

The Alzheimer’s Disease Sequencing Project (ADSP) is comprised of two Alzheimer’s Disease (AD) genetics consortia and three National Human Genome Research Institute (NHGRI) funded Large Scale Sequencing and Analysis Centers (LSAC). The two AD genetics consortia are the Alzheimer’s Disease Genetics Consortium (ADGC) funded by NIA (U01 AG032984), and the Cohorts for Heart and Aging Research in Genomic Epidemiology (CHARGE) funded by NIA (R01 AG033193), the National Heart, Lung, and Blood Institute (NHLBI), other National Institute of Health (NIH) institutes and other foreign governmental and non-governmental organizations. The Discovery Phase analysis of sequence data is supported through UF1AG047133 (to Drs. Schellenberg, Farrer, Pericak-Vance, Mayeux, and Haines); U01AG049505 to Dr. Seshadri; U01AG049506 to Dr. Boerwinkle; U01AG049507 to Dr. Wijsman; and U01AG049508 to Dr. Goate and the Discovery Extension Phase analysis is supported through U01AG052411 to Dr. Goate, U01AG052410 to Dr. Pericak-Vance and U01 AG052409 to Drs. Seshadri and Fornage.

## Consent statement

The study used publicly available de-identified data and was conducted in accordance with the Declaration of Helsinki and local ethical regulations. The Arizona State University IRB determined that the proposed activity is not research involving human subjects as defined by DHHS and FDA regulations. IRB review and approval by Arizona State University is not required.

## Author contributions

K.C., T.W, E.M.R., and Q.W. contributed to the concept and design of the study. Q.W. acquired and C.W.S. analyzed the data. C.W.S. and Q.W. contributed to drafting the text and preparing the figures. All authors approved the final version of the manuscript.

## Code availability

The analysis code for this work is available at https://github.com/clairewsu/tas2r38-mgam-a10bf

## Supplementary Figure Legends

Supplementary Figure 1: Longitudinal changes in PET imaging biomarkers (amyloid and tau), with fitted lines stratified by different taster groups in ADNI. In addition to WGS data, additional genetic data were obtained from ADNI microarray profiles to increase sample size.

Supplementary Figure 2: Longitudinal changes in CDR sum of boxes, with fitted lines stratified by different taster groups in NACC. P values and effect sizes for all the models are reported at the bottom.

Supplementary Figure 3: MGAM expression split by rs10246939 (g1) alternative allele count and Braak staging from MSBB cohort for four different brain regions.

Supplementary Figure 4: MGAM expression split by rs10246939 (g1) alternative allele count and Braak staging from MAYO cohort for two different brain regions.

Supplementary Figure 5: MGAM inhibitors slow down cognitive decline in T2D patients in PSM groups.

A) Kaplan-Meier curve comparing dementia-free probability between ever users and never users of MGAM inhibitors in PSM groups at ratio=2. n = 38 x 3, P = 0.062. B) Longitudinal changes in clinical assessment (CDRSUM) with fitted lines stratified by drug user groups in A). Group difference P = 0.010. Inset is standardized coefficients for the LME model. Significant terms are colored in red. C) same as A) with ratio = 3, n = 38 x 4 – 1 due to exact match, P = 0.14. D) same as B with ratio = 3, P = 0.010.

